# An integrated computational, clinical, and functional framework for assessing *PTPN11* (SHP2) variant effects on ERK signaling and neural crest cell behavior in Noonan spectrum disorders

**DOI:** 10.64898/2026.07.09.26357683

**Authors:** Mario Rodríguez-Martín, Kenza Cheriet, Sandrine Adiba, Vanessa Ribes, María Isidoro-García, Jesus Lacal, Pablo Prieto-Matos

**Affiliations:** Laboratory of Functional Genetics of Rare Diseases, Department of Microbiology and Genetics, University of Salamanca (USAL), Salamanca, Spain; GIR of Biomedicine of Rare Diseases, University of Salamanca, Salamanca, Spain; Instituto de Investigación Biomédica de Salamanca (IBSAL), Salamanca, Spain; Université Paris Cité, CNRS, INSERM, Institut Jacques Monod, F-75013 Paris, France; Université Paris Cité, CNRS, Epigenetics and Cell Fate, F-75013 Paris, France; Clinical Biochemistry Department, Hospital Universitario de Salamanca, Salamanca, Spain; Clinical Rare Diseases Reference Unit DiERCyL, Castilla y León, Spain; Department of Medicine, University of Salamanca, Salamanca, Spain; Department of Pediatrics, Hospital Universitario de Salamanca, Salamanca, Spain; Department of Biomedical and Diagnostics Science, University of Salamanca, Salamanca, Spain

**Author notes:** These authors contributed equally to this work (Mario Rodríguez-Martín, Kenza Cheriet). **Correspondence:** Jesus Lacal.

**Keywords:** RASopathies, Noonan syndrome, Noonan syndrome with multiple lentigines, *PTPN11*, SHP2, Disease modeling, Neural crest cells, Chick embryos

## Abstract

Germline mutations in *PTPN11* cause Noonan syndrome (NS) and NS with multiple lentigines (NSML), yet how specific variants drive divergent clinical outcomes through distinct signaling and developmental mechanisms remains unclear. We find that germline and somatic mutations converge on N-SH2 and PTP domains but diverge at residue-level hotspots, reflecting distinct selective pressures. Clinical stratification of 18 pediatric patients reveals four distinct phenotypic classes including (i) the NSML-associated c.1403C>T (T468M) variant, characterized by lentigines, moderate growth impairment, and distinctive facial features; (ii) variants including the VUS c.1282G>A (V428M) and c.1432A>G (I478V), which were associated with cognitive deficits and variable growth impairment; (iii) c.1471C>A (P491T) and c.1472C>T (P491L), predominantly affecting cardiac and growth phenotypes with limited neurocognitive features; and (iv) a severe, multisystem class comprising c.172A>G (N58D), c.178G>A (G60S), c.844A>G (I282V), c.922A>G (N308D), and c.923A>G (N308S), spanning cardiac, growth, cognitive, and craniofacial abnormalities. Biochemical profiling in HEK293T cells revealed that *PTPN11* variants stratify beyond simple gain/loss-of-function dichotomies into strong ERK-dependent hyperactivation, moderate ERK activation with variable protein stability and the paradoxical c.1282G>A variant, which did not increase ERK phosphorylation. In vivo, this variant drove excessive neural crest cell migration in chick embryos, suggesting that its effects on NCC migration may involve ERK-independent mechanisms or context-dependent signaling not captured by steady-state assays. ERK activation did not strictly correlate with clinical severity, yet these functional differences were associated with distinct growth, cardiac, pigmentation, and neurodevelopmental outcomes. Our data suggest lineage-specific sensitivity to SHP2 dosage, with dorsal root ganglia neurons appearing more vulnerable to reduced SHP2 stability than melanocyte precursors. Although direct correlations between specific signaling defects and individual clinical features remain complex, our findings provide a refined framework for *PTPN11* variant classification, and reveal unexpected SHP2 functions in neural crest development.

## Introduction

The functional interpretation of genetic variants remains a major bottleneck in human genetics. While the most common pathogenic variants have been extensively characterized, the majority of variants identified in clinical settings, particularly variants of uncertain significance (VUS), remain difficult to link to phenotypic outcomes ^1,2^. Here, we address this challenge by focusing on *PTPN11*, which encodes SHP2, a nonreceptor protein tyrosine phosphatase that plays a central role in cellular signaling. SHP2 functions as a key signaling hub downstream of growth factor, cytokine, and integrin receptors, coordinating multiple pathways, most prominently the RAS/ERK cascade, as well as PI3K/AKT, JAK/STAT, NF-κB, and β-catenin signaling ^3,4^.

Somatic mutations in *PTPN11* are frequently identified in hematological malignancies, such as juvenile myelomonocytic leukemia (JMML), acute myeloid leukemia (AML), and B-cell acute lymphoblastic leukemia (B-ALL), as well as in solid tumors including melanoma and glioblastoma ^5,6^. In parallel, germline mutations in *PTPN11* underlie two RASopathies, including Noonan syndrome (NS) and Noonan syndrome with multiple lentigines (NSML), a group of neurodevelopmental disorders associated with altered RAS pathway signaling ^7–11^. These autosomal dominant disorders display overlapping phenotypes including craniofacial abnormalities, congenital heart defects, and growth delay, while NSML is further distinguished by multiple lentigines, sensorineural hearing loss, and ocular hypertelorism ^7,9,12–15^. Importantly, structural and functional studies consistently show that somatic mutations drive strong gain-of-function effects on SHP2 phosphatase activity, whereas the consequences of germline variants associated with NS and NSML are more complex and extend beyond a simple activity-based framework ^5,6,16^.

The most extensively studied *PTPN11* mutations in NS and NSML, as in cancer, affect residues within two key domains that regulate SHP2 activity, the N-terminal SH2 domain (N-SH2) and the catalytic protein tyrosine phosphatase (PTP) domain. These domains are functionally coupled through an autoinhibitory mechanism, whereby, under basal conditions, SHP2 adopts a conformation in which the N-SH2 domain occludes the catalytic site of the PTP domain ^17,18^. This inhibition is relieved upon activation of upstream receptors, such as RTKs, as binding to phosphotyrosine-containing motifs induces a conformational change toward an open and active state ^19,20^. The classical model posits that NS-associated mutations, similarly to cancer-associated variants, affect residues within the N-SH2 domain (e.g. D61, A72, E76) or the PTP domain (e.g. G503) and destabilize the autoinhibited conformation, thereby promoting increased SHP2 activity ^16,21–23^; albeit with effects that are milder than those observed for cancer-associated mutations ^24^. In contrast, NSML-associated mutations, which preferentially affect residues within the PTP domain (e.g. Y279, T468), have been shown to decrease or abolish SHP2 phosphatase activity ^16,25–27^.

However, this binary gain- versus loss-of-function model of NS and NSML is increasingly challenged. The phenotypic overlap between NS and NSML, the identification of NSML-associated variants exhibiting gain-of-function properties in specific signaling contexts, the presence of variants associated with both conditions, and the involvement of mutations affecting additional residues all point to a more complex functional landscape. For instance, several NSML-associated mutations, although catalytically impaired, have been shown to promote sustained activation of downstream pathways such as AKT/mTOR signaling ^26,28,29^. Conversely, certain NS-associated variants display context-dependent or moderate effects on phosphatase activity, further blurring the distinction between classical gain- and loss-of-function categories ^16,30^. In addition, overlapping or closely positioned residues within the PTP domain have been reported to give rise to either NS or NSML phenotypes depending on the specific substitution, highlighting the importance of subtle structural and conformational effects ^25,26^. Furthermore, emerging evidence indicates that SHP2 exerts functions beyond its catalytic activity, including scaffolding roles and the regulation of protein–protein interactions, suggesting that the impact of disease-associated variants cannot be fully explained by changes in phosphatase activity alone ^17^.

This complexity is further compounded by strong cell type-dependent effects. Studies in model organisms have shown that *PTPN11* variants impact multiple embryonic lineages, with particularly pronounced effects in neural crest cells (NCCs), a highly migratory population that gives rise to craniofacial structures, melanocytes, and components of the peripheral nervous system ^17,19,31,32^. Consistent with this, the effects of *PTPN11* variants differ markedly across tissues. For example, NS-associated mutations such as D61G and Q79R promote hyperactivation of ERK signaling across tissues; however, in cardiac lineages they drive increased proliferation, contributing to hypertrophic cardiomyopathy, whereas in NCCs they primarily disrupt migration and survival, leading to craniofacial defects ^31–33^. Similarly, NSML-associated mutations can mimic loss-of-function effects in some settings, such as reduced growth, while producing distinct phenotypes in NCC-derived ^17^. At the mechanistic level, these differences reflect both phosphatase-dependent and -independent functions of SHP2, including roles mediated by its C-SH2 domains in regulating p53-dependent apoptosis in the brain and neural crest ^34^.

Building on these observations, we developed a combinatorial approach to systematically investigate genotype–phenotype relationships of germline *PTPN11* variants associated with NS/NSML. To this end, we integrated computational analysis of all reported variants to identify regions of SHP2 prone to NS/NSML-associated alterations, deep clinical phenotyping of a pediatric NS/NSML cohort, and biochemical characterization of selected variants in cellular systems. In addition, we leveraged an accessible *in vivo* model, the chick neural tube, to functionally interrogate variant effects within a neural crest context. This strategy uncovers distinct functional classes of variants that extend beyond a simple gain- or loss-of-function framework and demonstrates that neural crest cells are particularly sensitive to variant-specific perturbations of SHP2 activity.

## Results

### *PTPN11* mutational hotspots converge on N-SH2/PTP domains but diverge between germline and somatic contexts

To define the landscape of *PTPN11* variation across development and cancer, we compared the distribution of germline variants (ClinVar) and somatic mutations (COSMIC). This analysis aimed to identify functionally constrained domains and to assess whether distinct selective pressures shape *PTPN11*mutations in inherited versus acquired disease contexts.

We first analyzed the germline landscape using 769 *PTPN11* coding variants from ClinVar. These variants included missense, synonymous, and truncating changes (Figure 1A). Recurrent hotspots (each ≤1% of cases, ≥7 occurrences) were found at residues N58, G60, D61, E76, F285, T468, G503, and Q510 (Figure 1A). Among these 769 variants, 180 (23.4%) were classified as pathogenic or likely pathogenic, 203 (26.4%) as benign or likely benign, 340 (44.2%) as VUS, 44 (5.7%) as conflicting and 2 (0.3%) had no classification (Figure 1B). As expected, the analysis of gnomAD allele frequencies confirmed that benign variants exhibited significantly higher population frequencies than pathogenic ones (Figure 1C). Pathogenic variants were unevenly distributed along SHP2, enriched in the N-SH2 and PTP domains but underrepresented in the C-SH2 domain and interdomain regions (Figure 1D). Missense variants clustered predominantly between residues N58–E76 within the N-SH2 domain and at the C-terminal portion of the PTP domain (around residues P491 and Q510), with additional clusters in the N-terminal PTP domain and, to a lesser extent, the C-SH2 domain (Figure 1E). Clinically, these 180 pathogenic or likely pathogenic variants were associated with RASopathy-related conditions (62), NS (47), NSML (14), metachondromatosis (14), cardiovascular disorders (2), male infertility (2), short stature (2), autism spectrum disorder (ASD) (1), and juvenile myelomonocytic leukemia (JMML) (1), while 35 remained unclassified (Figure S1A). Interestingly, 48% of the variants were associated with multiple phenotypes, indicating substantial pleiotropy.

**Figure 1.**
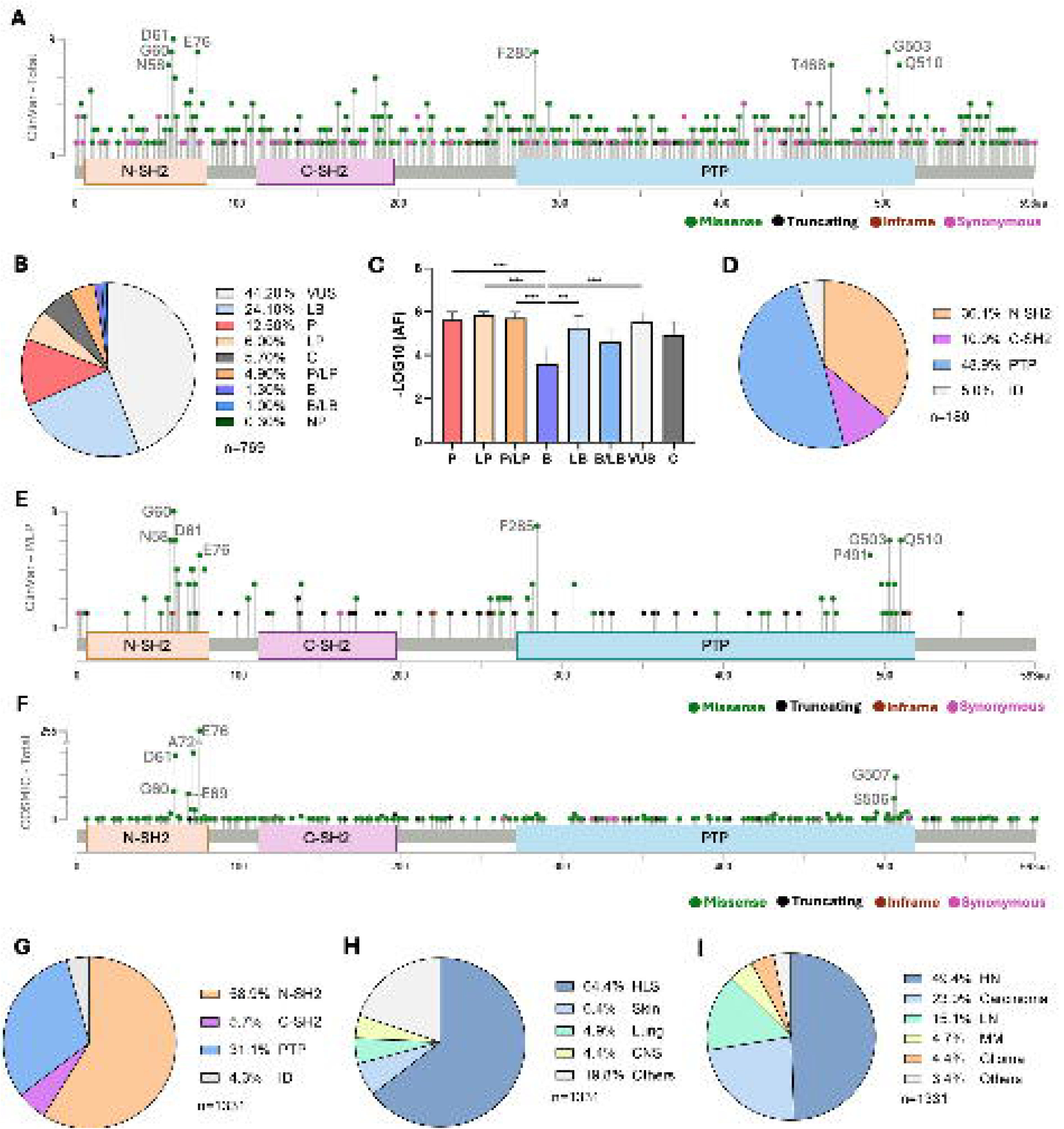
Landscape of germline and somatic *PTPN11* variants. (A) Lollipop diagram of ClinVar germline variants shows broad distribution across N-SH2 and PTP domains. (B) Pie chart of pathogenicity classifications. (C) gnomAD allele frequencies correlate with pathogenicity, benign variants show higher frequencies than pathogenic ones. (D) Domain distribution of pathogenic germline variants. (E) Lollipop diagram of pathogenic missense variants. (F) Lollipop diagram of pathogenic somatic COSMIC mutations. (G) Domain distribution of pathogenic somatic mutations annotated in COMIC. (H) Primary tissue distribution, and (I) histological classification. P: pathogenic; LP: likely pathogenic; VUS: variant of uncertain significance; B: benign; NR: not reported; C: conflicting interpretation; ID: inter domain region; HLS: hematopoietic and lymphoid systems; CNS: central nervous system; HN: hematopoietic neoplasm; C: carcinoma; LN: Lymphoid neoplasm; MM: malignant melanoma.

The somatic mutational landscape of *PTPN11*, comprising 1331 coding mutations reported as pathogenic in COSMIC, shows that mutations cluster prominently within the N-SH2 and PTP domains, due to recurrent hotspots including G60, D61, E69, A72, and E76 in the N-SH2 domain, and S506 and G507 in the PTP domain (Figure 1F). Quantification by domain shows that the N-SH2 accounts for 58.9% of somatic mutations, the PTP for 31.1%, whereas the C-SH2 (5.7%) and interdomain regions (4.3%) are markedly underrepresented (Figure 1G). These 1331 *PTPN11* mutations were distributed across 27 primary tissue types, with the highest frequencies in hematopoietic and lymphoid tissues (64.4%), followed by skin (6.4%), lung (4.9%) and central nervous system (4.4%) (Figure 1H). At the histological level, mutations were enriched in hematopoietic neoplasms (49.4%), carcinomas (23%), and lymphoid neoplasms (15.1%), with lower frequencies in melanomas (4.7%) and gliomas (4.4%) (Figure 1I). Accordingly, *PTPN11* mutations were most observed in JMML, acute myeloid leukemia (AML), adenocarcinoma (ADC), B cell acute lymphoblastic leukemia (B-ALL), desmoplastic melanoma (DM), and glioblastoma (GBM) (Figure S1B).

### Genotype-phenotype mapping of *PTPN11* variants in pediatric Noonan spectrum disorders reveals mutation-specific effects on growth, cardiac defects, and neurodevelopment

To investigate genotype-phenotype correlations in *PTPN11*-related Noonan spectrum disorders, we first performed a comprehensive clinical characterization of 18 pediatric patients. All patients met established diagnostic criteria for either Noonan syndrome (NS, n=14) as defined by van der Burgt et al. ^35^ or Noonan syndrome with multiple lentigines (NSML, n=4) as defined by Gelb and Tartaglia. ^36^. The phenotypic assessment focused on neurodevelopment, cardiac involvement, growth, and facial dysmorphism (Table 1). WES and targeted analysis of RASopathy-associated genes identified 10 distinct heterozygous *PTPN11* missense variants, including c.172A>G (p.N58D) and c.178G>A (p.G60S) in the N-SH2 domain, and eight additional variants mapping to the PTP domain, c.844A>G (p.I282V), c.922A>G (p.N308D), c.923A>G (p.N308S), c.1282G>A (p.V428M), c.1403C>T (p.T468M), c.1432A>G (p.I478V), c.1471C>A (p.P491T), and c.1472C>T (p.P491L) (Figure 2A; Table 1). To assess whether the identified *PTPN11* variants might affect pre-mRNA splicing, we performed SpliceAI predictions. All variants yielded scores below the established splicing effect threshold of 0.2 (range 0.00–0.09), indicating that none are predicted to alter splicing.

**Figure 2.**
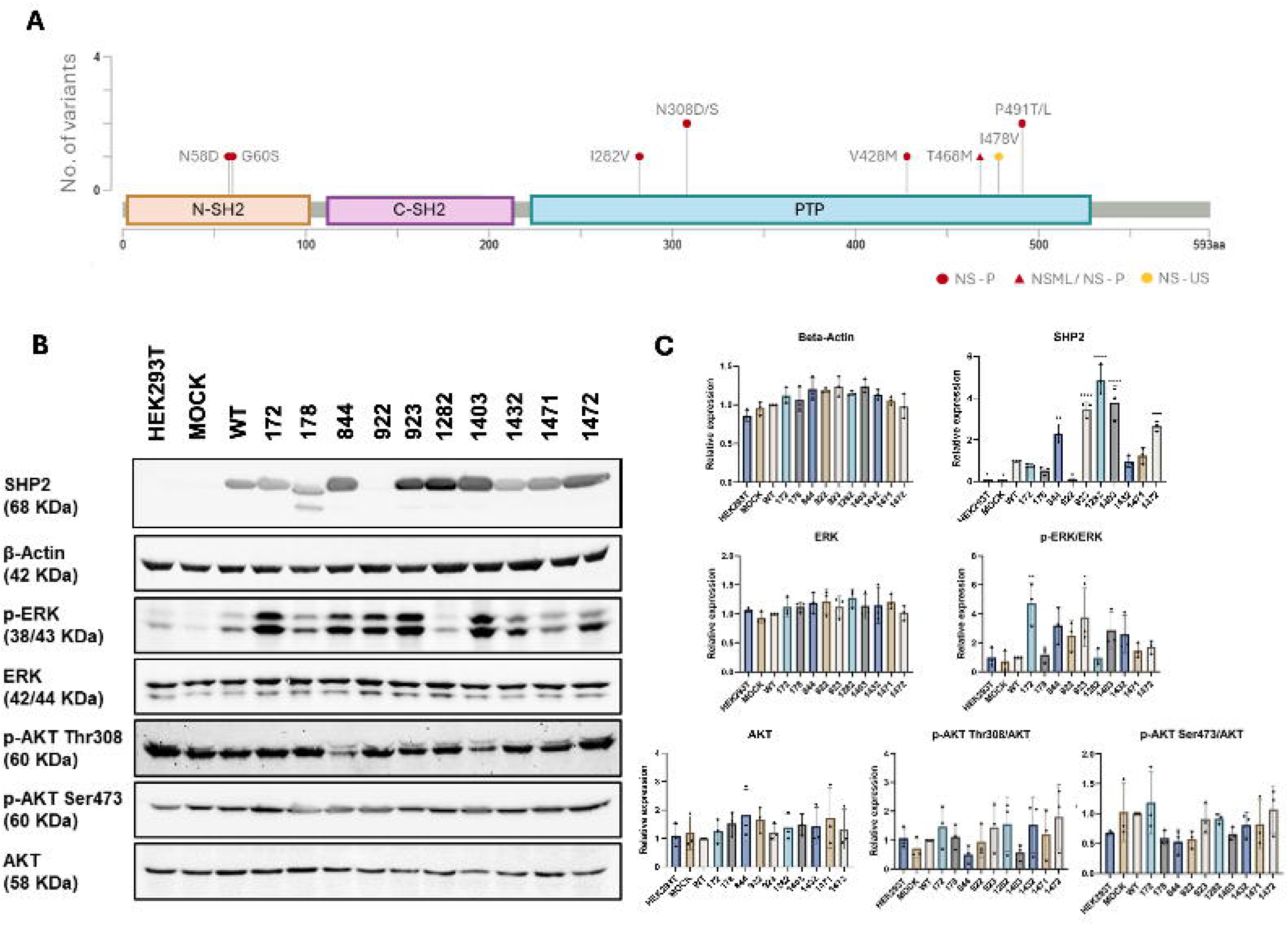
Functional characterization of *PTPN11* variants identified by our clinical genetics unit. **(A)** Schematic representation of the ten pediatric variants detected in patients with NS and NSML along the SHP2 protein. **(B)** Western blot analysis of stable HEK293T cell lines expressing *PTPN11* wild-type (WT) or variant constructs, alongside control conditions. Representative immunoblots show the expression of SHP2, as well as downstream signaling components including ERK, phosphorylated ERK (pERK), AKT, and phosphorylated AKT at Ser473 and Thr308. β-actin was used as a loading control to confirm equal protein input. **(C)** Quantification of immunoblots shown in (B). Mean ± SD.

**Table 1.**
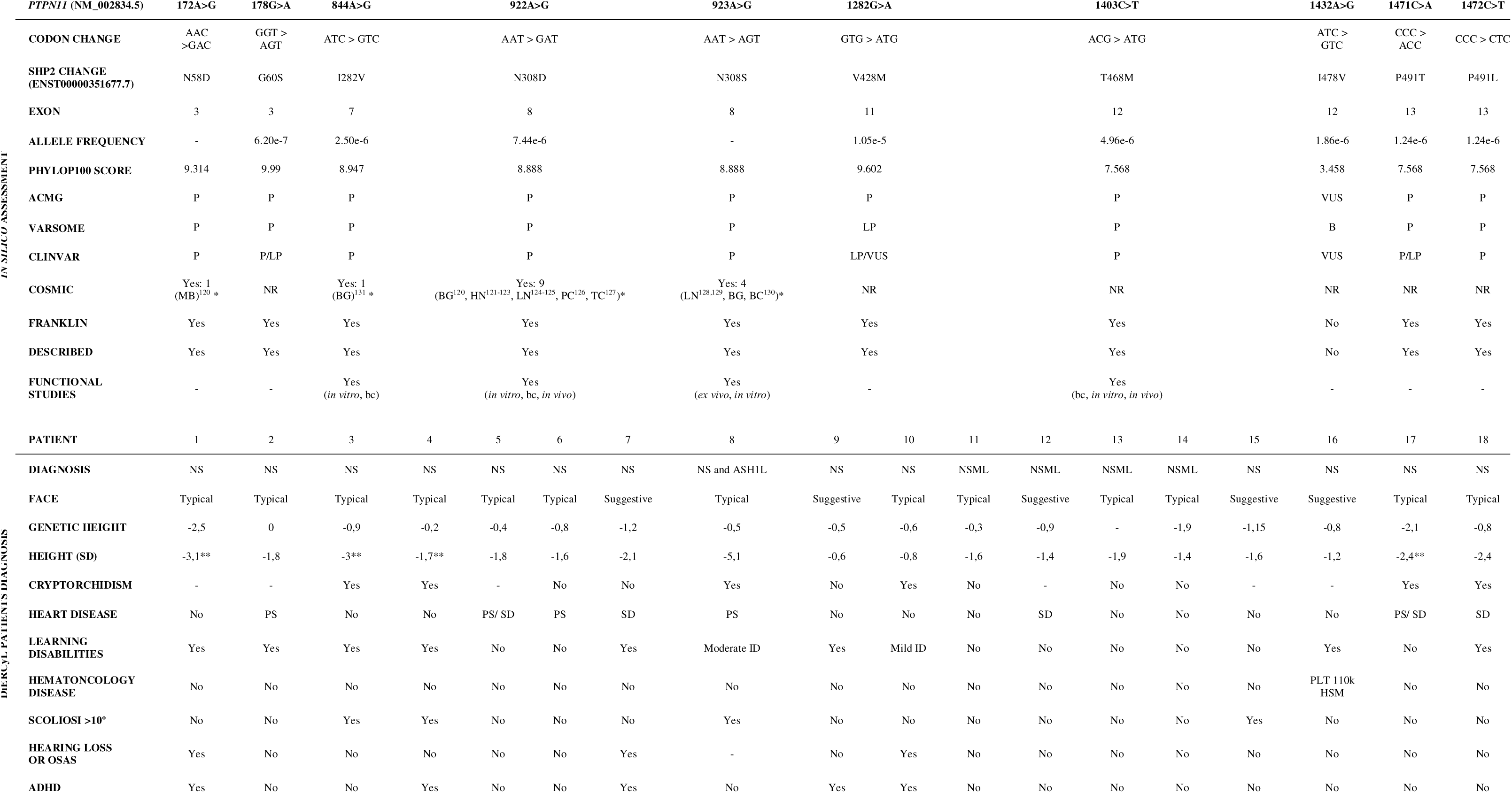
Clinical and genetic data of 18 pediatric patients harboring ten *PTPN11* variants cohort. Codon and residue changes, exon in which they are located, allele frequency by gnomeAD, evolutionary conservation by PhyloP100, predicted pathogenicity based on ACMG, Varsome and ClinVar annotations, associations with various cancers as reported in the COSMIC database, prevalence gathered in the Franklin dataset, and functional studies reported in the literature are shown for each variant in the upper section of the table. The lower part of the table details the clinical phenotypes of the 18 patients from the DiERCyL cohort. P: pathogenic; LP: likely pathogenic; VUS: variant of uncertain significance; B: benign; NR: not reported; PNET-MB: Primitive neuroectodermal tumor-medulloblastoma; BG: brain glioma; HN: hematopoietic neoplasm; LN: lymphoid neoplasm; PC: prostate carcinoma; TC: thyroid carcinoma; BC: breast carcinoma; EC: esophagus carcinoma. bc: biochemical. *Found in NM_001330437.2 (SHP2 isoform of 597aa - p.407insQALL), but at until residue 408 the coding sequence is the same. NS: Noonan syndrome. NSML: Noonan syndrome with multiple lentigines. **: Growth hormone treatment. PS: Pulmonary stenosis. SD: Septal defect. ID: Intellectual disability. PLT: platelet count. HSM: hepatosplenomegaly. OSAS: Obstructive sleep apnea syndrome. ADHD: Attention Deficit Hyperactivity Disorder. ASH1L: pathogenic variant in *ASH1L*:c.708dupA.

All variants were classified as pathogenic or likely pathogenic by ACMG guidelines, with the exception of c.1282G>A (V428M) and c.1432A>G (p.I478V), which were classified as a VUS. Most variants affected highly conserved residues (PhyloP scores >7.5), except c.1432A>G (3.458), although this remains above the threshold indicative of evolutionary constraint (2.27). All identified *PTPN11* variants, except c.1432A>G (only in ClinVar), have been previously reported in the literature and public databases^16,37,38^. All variants exhibited very low allele frequencies in gnomAD (1.24×10LL to 1.05×10LL), consistent with their rare, disease-associated nature. Functional studies have been previously reported for c.844A>G, c.922A>G, c.923A>G, and c.1403C>T, demonstrating gain-of-function effects in vitro and in vivo ^23,32,39–41^. De novo inheritance occurred in 61% of patients (11/18) and was associated with earlier diagnosis (mean age 6.2 years) and more frequent cardiac involvement (82%, 9/11), whereas inherited variants (6/18, all but one maternal) presented with milder phenotypes and later diagnosis (mean age 9.1 years).

The four unrelated patients with a molecular diagnosis of NSML (patients 11–14) all carried the same c.1403C>T variant. Clinically, NSML patients were uniformly distinguished from NS patients by the presence of lentigines, and the complete absence of pulmonary stenosis (PS) (0% vs. 38% in NS) and absence of learning disabilities (0% vs. 77% in NS). Septal defects (SD) were present in 25% of NSML patients (1/4) and 29% of NS patients (4/14). These findings establish T468M as a potential genetic marker for NSML as all four unrelated NSML patients carried this variant. This is consistent with previous studies identifying T468M as a NSML-associated mutation that, in mouse models, confers leanness and protects against diet-induced obesity ^42^.

Pulmonary stenosis (PS) was associated with c.178G>A (G60S), c.922A>G (N308D), c.923A>G (N308S) and c.1471C>A (P491T), whereas septal defects (SD) were found in patients harboring c.922A>G, c.1403C>T (T468M), c.1471C>A and c.1472C>T (P491L). No cardiac defects were observed in the patients carrying c.172A>G (N58D), c.1282G>A (V428M) or c.1432A>G (I478V). Marked short stature (height SDS ≤ –2) was associated with c.172A>G, c.844A>G (I282V), c.1471C>A and c.1472C>T. The most severe growth retardation (SDS –5.1) was observed in a patient carrying c.923A>G (patient 8, who also carried an *ASH1L* variant). The clinical features of this patient cannot be unambiguously attributed to *PTPN11* due to the co-occurring *ASH1L* variant, which raises the possibility of a modifying or synergistic effect, though confirmation in larger cohorts is required. Patients with the NSML-associated variant c.1403C>T exhibited milder growth deficits (height SDS range –1.4 to –1.9), whereas those with c.1432A>G and c.1282G>A showed only mild or no growth impairment. Learning disabilities (LD) were predominantly associated with NS-related variants, particularly c.178G>A, c.844A>G, c.1432A>G and c.1472C>T. Combined LD and attention deficit hyperactivity disorder (ADHD) were observed with c.172A>G, c.922A>G (two of four cases) and c.1282G>A. By contrast, the NSML-associated variant c.1403C>T was consistently associated with preserved cognitive function in all four patients. Across the cohort, LD was diagnosed in three of six females (50%) and six of twelve males (50%), whereas ADHD was found in four of twelve males (33%) and one of six of females (17%). Typical NS facial features, including broad forehead, down-slanting palpebral fissures, deep nasal bridge, were observed in all patients carrying NS-associated variants. Cryptorchidism was present in six of twelve male patients (50%) and occurred exclusively in NS. Scoliosis >10° was observed in four of fourteen NS patients (29%) but in no NSML patients. Hearing loss or obstructive sleep apnea occurred in three of fourteen NS patients (21%). Interestingly, hematological findings were limited to the c.1432A>G VUS, with thrombocytopenia (platelet count 110 × 10L/L) and hepatosplenomegaly; no malignancies were reported in the cohort. There is no evidence of sex bias or age dependence in the clinical manifestations of this cohort. The phenotypic variability is driven by genotype (specific *PTPN11* variant) rather than sex or age.

### *PTPN11* variants differentially affect SHP2 protein stability and ERK activation, with most driving hyperactivation and one impairing signaling

To dissect the molecular mechanisms of *PTPN11* variants, we stably expressed wild-type and mutant proteins in HEK293T cells via lentiviral transduction. For each construct, transduced cells were pooled to avoid clonal selection biases, and Western blot analyses were performed from three independent transduction experiments. We then evaluated SHP2 protein abundance and activation of its canonical downstream effectors, ERK and AKT, compared with parental and empty-vector controls. Western blot results confirmed the overexpression of SHP2 in most samples (Figures 2B, C). Variants c.172A>G, c.178G>A, c.1432A>G, and c.1471C>A showed expression levels comparable to wild-type, whereas the remaining variants displayed a 2- to 5-fold increase in SHP2 abundance (Figure 2C), suggesting increased protein stability. Interestingly, the c.178G>A (G60S) variant exhibited altered electrophoretic mobility, appearing as a double band (Figure 2B), suggesting increased partial proteolytic degradation. The low levels of the c.922A>G (N308D) variant suggested impaired protein stability, potentially due to misfolding or accelerated proteasomal degradation. Nevertheless, this variant drove a reproducible 2.5-fold increase in ERK phosphorylation compared with wild-type controls (Figure 2C). β-actin levels were consistent across all samples, supporting reliable normalization of the quantitations (Figure 2B, 2C).

ERK phosphorylation (Thr202/Tyr204) was enhanced in the majority of *PTPN11* variants relative to wild-type SHP2. Six of the ten variants including c.172A>G, c.844A>G, c.922A>G, c.923A>G, c.1403C>T, and c.1432A>G, drove 2- to 5-fold increases in pERK levels. This ERK hyperactivation was not driven by increased SHP2 protein expression but to the type of mutation in SHP2. The c.922A>G (N308D) variant, exhibited very low SHP2 abundance yet produced 2.5-fold elevated pERK, while c.1282G>A (V428M) showed the highest SHP2 levels but no increase in pERK. These data suggest that the observed pathway activation is primarily dirven by mutation-intrinsic effects on SHP2 activity in this celullar context (Figure 2B, 2C). Variants c.172A>G (N58D) and c.923A>G (N308S) drove the strongest ERK hyperactivation, 5-fold and 4-fold over WT, respectively (p < 0.01 and p < 0.05). Variants c.844A>G (I282V), c.922A>G (N308D), c.1403C>T (T468M), and c.1432A>G (I478V) showed intermediate ERK activation (2- to 3-fold over WT). Variants c.178G>A (G60S), c.1282G>A (V428M), c.1471C>A (P491T), and c.1472C>T (P491L) showed pERK levels comparable to wild type. No variant led to a significant reduction in pERK below wild-type levels. To determine whether SHP2 variants also affect the PI3K/AKT pathway, we assessed AKT phosphorylation at Thr308 (PDK1 site) and Ser473 (mTORC2 site). Overall, AKT phosphorylation levels showed limited variability across variants and did not reveal any differences relative to wild-type or control conditions (Figure 2B, 2C). These results indicate that, in this cellular system, *PTPN11* variants predominantly impact the MAPK pathway rather than the PI3K/AKT axis.

### Embryonic neurogenic lineages represent a permissive context for the expression of *PTPN11* variants

To further investigate the developmental consequences of *PTPN11* variants *in vivo*, we expressed representative NS-associated variants displaying distinct biochemical profiles in the neural tube of chick embryos. Electroporations were performed at Hamburger-Hamilton (HH) stages 10-11, a developmental stage at which the neural tube contains both spinal cord neural progenitors and neural crest cells (NCCs), two cell populations frequently affected in RASopathies (Figure 3A). Wild-type *PTPN11* and three representative variants (c.172A>G, c.922A>G and c.1282G>A) were cloned into the pCIG vector and unilaterally electroporated into the neural tube. As a control for SHP2 overexpression effects, a pCIG-empty vector was electroporated in parallel in all experiments. Embryos were analyzed 24 and 48 hours post-electroporation (hpe). As expected, GFP-positive cells were detected within the neural tube at both stages and, at 48 hpe, in NCC-derived tissues including dorsal root ganglia (DRG), melanocyte precursors (MPs), and Schwann cell precursors (SCPs) (Figure 3A,B). We first examined SHP2 expression levels by immunostaining. At both 24 and 48 hpe, SHP2 levels in electroporated cells were generally higher than endogenous levels, confirming efficient transgene expression (Figure 3B). Notably, all mutant variants tended to produce higher SHP2 levels than wild-type *PTPN11*, particularly at 48 hpe, consistent with increased protein stability. SHP2 displayed the expected cytoplasmic localization in both neural progenitors and NCC derivatives. Together, these observations indicate that NS-associated *PTPN11* variants are robustly expressed in embryonic neurogenic lineages and suggest that several mutations may enhance SHP2 protein stability *in vivo*.

**Figure 3.**
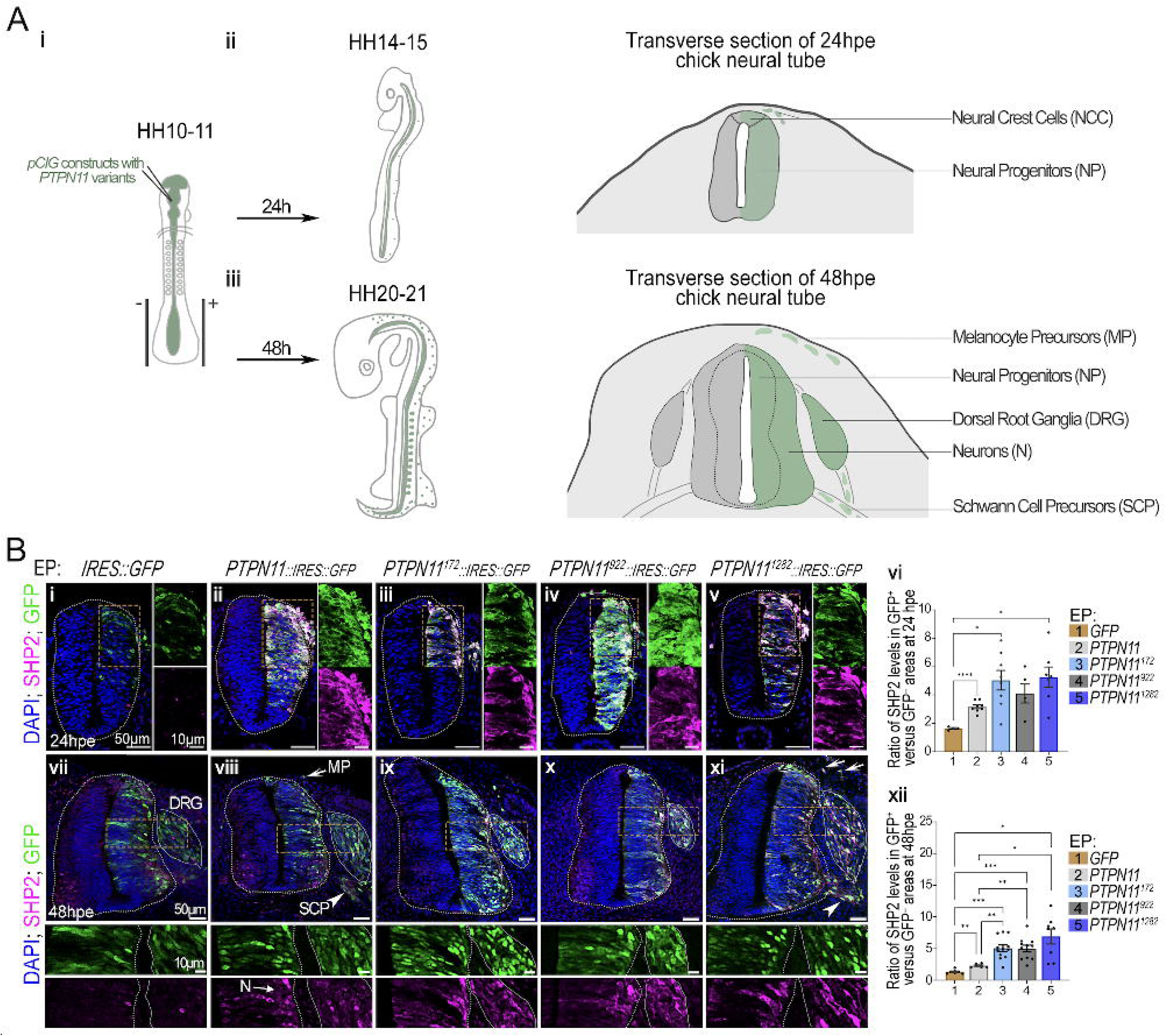
Quantitative analysis of SHP2 WT and mutant variant expression in the chick neural tube. **(A)** (i) Schematic representation of chick embryos at the time point of plasmid injection and electroporation, HH10-11. (ii) Upper panel showing schematic of a transverse section of the chick neural tube at 24 hpe at the brachial level, illustrating neural progenitors in the neural tube as well as NCC located in the most dorsal part of the neural, with the migrating ones located outside of the neural tube. (iii) Likewise at 48 hpe including dorsal subepidermal melanocyte precursors (MP), the dorsal root ganglia (DRG), and Schwann cell precursors (SCP) along the ventral root nerve. **(B)** (i–v; vii–xi) Immuno-labelling for SHP2 (magenta), GFP (green), and DAPI (blue) on transverse sections of chick embryos at 24 and 48 hpe with the indicated constructs. Orange dotted squares in overview panels indicate the regions shown at higher magnification; dotted lines delineate the neural tube, and thin lines indicate the dorsal root ganglia (DRG). Melanocyte precursors (MP) are indicated by white arrows, and Schwann cell precursors (SCP) by white arrowheads. Scale bars: 50 µm (overview panels) and 10 µm (higher-magnification panels). (vi, xii) Quantification of the ratio of SHP2 levels in GFPL cells relative to GFPL cells at 24 hpe (vi) and 48 hpe (xii) with the indicated constructs (n > 3 embryos per condition; dots represent individual embryos; bars indicate mean ± s.e.m.; Dunnett’s T3 multiple comparisons test: *p < 0.05; **p < 0.01; ***p < 0.001; ****p < 0.0001).

### *PTPN11* variants alter neural crest cell allocation without affecting neurogenic differentiation

We next investigated whether *PTPN11* variants affect neural and neural crest differentiation. Immunostaining for SOX1, SOX10 and HuC/D at 48 hpe revealed the expected distribution of neural progenitors, neural crest-derived progenitors and differentiated neurons in all conditions (Figure 4A,B). Quantification of GFP^+^SOX1^+^ and GFP^+^HuC/D^+^ areas within the neural tube, and calculation of the SOX1/HuC/D area ratio, did not reveal significant differences between conditions. Similarly, the GFP^+^HuC/D^+^ area within dorsal root ganglia (DRG) was unchanged upon expression of *PTPN11* variants (Figure 4C). Together, these results indicate that *PTPN11* variants do not markedly affect spinal cord or DRG neurogenesis at this stage.

**Figure 4.**
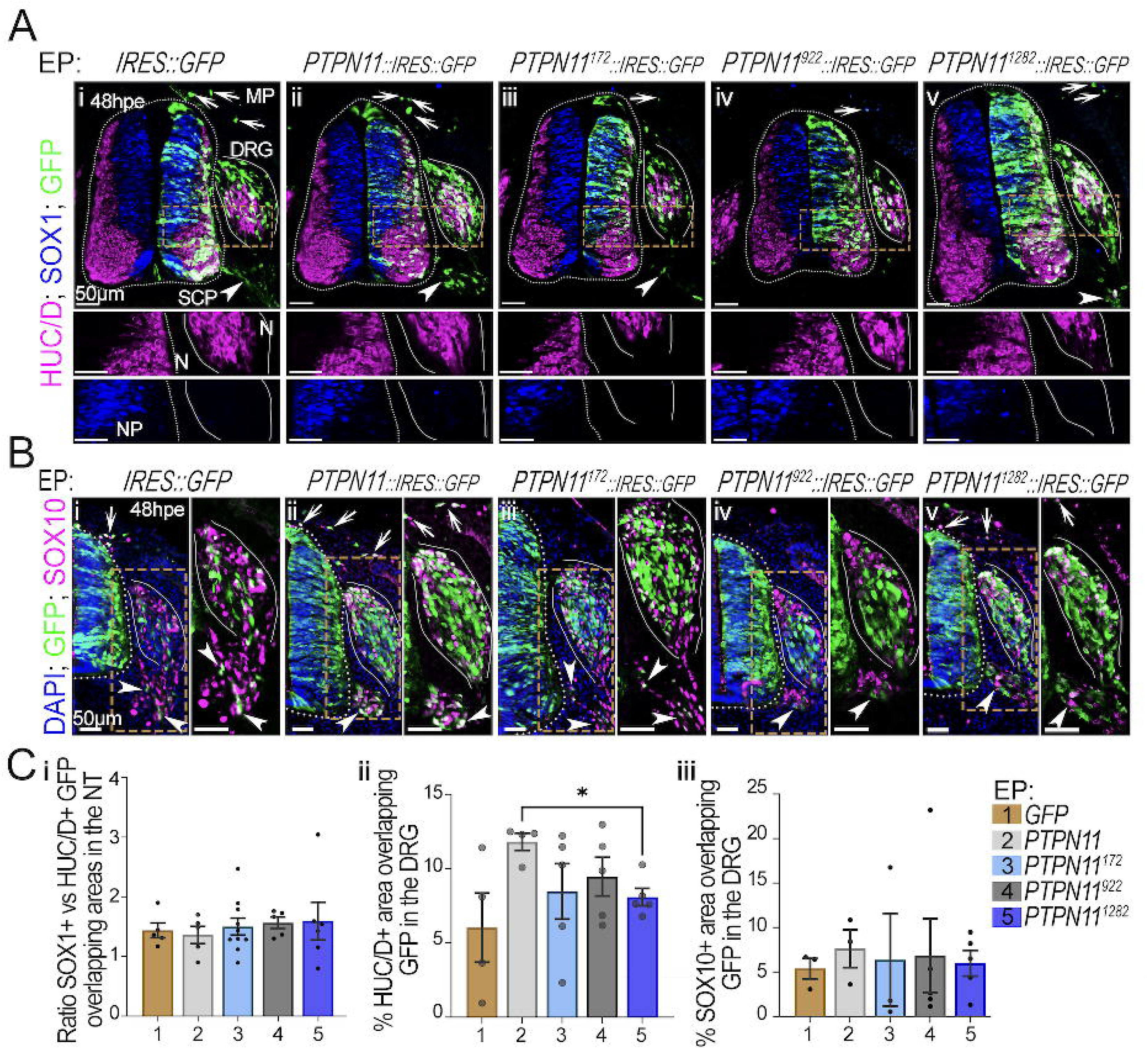
Neuronal differentiation potential of *PTPN11* WT- or variant-expressing cells in histological of chick embryos. **(A)** Immunolabeling for HuC/D (magenta), SOX1 (blue), and GFP (green) on transverse sections of chick embryos at 48 hpe with the indicated constructs. Orange dotted boxes in panels (i–v) indicate two regions shown at higher magnification below. Dotted lines delineate the neural tube, and thin lines, the DRG. White arrows: melanocyte precursors (MP), white arrowheads: Schwann cell precursors (SCP). Scale bars: 50 µm. **(B)** Immunolabeling for SOX10 (magenta), GFP (green), and DAPI (blue) on transverse sections of chick embryos at 48 hpe with the indicated constructs. Each of the right panels is a higher-magnification view of the regions indicated by orange dotted boxes in the left panels. Dotted lines delineate the neural tube, and thin lines delineate the DRG. White arrows point at MP, and white arrowheads at SCP. Scale bars: 50 µm. **(C)** Ratio of SOX1L vs HuC/D^+^ areas overlapping with GFP areas in the neural tube (i), and SOX10 signal overlapping with GFP in the DRG (ii), quantified in transverse sections at 48 hpe. Dots: value per section; bars: mean ± s.e.m. Statistical analysis was performed on n ≥ 3 embryos per condition, using Tukey’s multiple comparisons test for (i) and unpaired Mann-Whitney tests between each condition for (ii) (*P < 0.05, **P < 0.01, ***P < 0.001, ****P < 0.0001).

While analyzing SHP2 expression and lineage markers, we noticed differences in the distribution of GFP-positive neural crest derivatives between *PTPN11* variants (Figures 4-5). To characterize these effects, we performed morphometric analyses on transverse sections of chick embryos at 48 hpe. DRG area and elongation were measured as descriptors of ganglion morphology, while SCP clusters along peripheral axons and GFP-positive cells within the dorsal subcutaneous mesenchyme were quantified on each section (Figure 5A,B). Upon expression of *PTPN11*^922^, DRGs were smaller and displayed a more elongated morphology. In contrast, *PTPN11*^1282^ primarily affected DRG shape, resulting in more elongated ganglia without significantly altering their size. WT *PTPN11* and *PTPN11*^172^ had little effect on these parameters. Furthermore, expression of WT *PTPN11* and *PTPN11*^172^ and *PTPN11*^1282^ increased the number of SCP clusters detected along ventral roots, whereas *PTPN11*^922^ did not. Only *PTPN11*^1282^ additionally increased the number of GFP-positive cells within the dorsal subcutaneous domain. To confirm and extend these observations, we analyzed whole-mount embryos immunostained for GFP and ISLET1/2, allowing visualization of electroporated NCC derivatives and DRG neurons along the entire anterior-posterior axis (Figure 5C,D, S1). This approach enabled the simultaneous examination of brachial, thoracic and lumbosacral regions and provided a global view of DRG organization, peripheral nerves and melanocyte precursor distribution through 3D confocal reconstructions. Consistent with observations from transverse sections, DRG appeared smaller in embryos expressing *PTPN11*^922^ and *PTPN11*^1282^ throughout the anterior-posterior axis (Figure 5C). In addition, SCPs were noticeably more abundant along peripheral nerves in WT *PTPN11*, *PTPN11*^172^ and *PTPN11*^1282^ embryos than in control embryos (Figure 5C). In WT *PTPN11* embryos, SCPs were primarily detected in the distal portions of motor nerves, whereas in *PTPN11*^172^ and *PTPN11*^1282^ embryos they were distributed along much of the nerve trajectory from proximal to distal regions. In contrast, *PTPN11*^922^ embryos displayed only a few SCPs and more closely resembled the control condition (Figure 5C). Additional projections at tail levels (Figure 5D) and in the skin at thoracic levels (Figure S1) revealed melanocyte precursor populations. Upon WT *PTPN11* expression, these cells appeared relatively evenly distributed throughout the skin. In contrast, all mutant *PTPN11* variants induced a clustered distribution of melanocyte precursors. These clusters occupied broader skin territories in *PTPN11*^172^ and *PTPN11*^1282^ embryos, whereas fewer melanocyte precursors were observed in *PTPN11*^922^ embryos. Thus, *PTPN11* variants differentially affect the allocation and spatial organization of neural crest derivatives, generating distinct developmental phenotypes. These findings highlight the sensitivity of neural crest development to variant-specific perturbations of SHP2 function.

**Figure 5.**
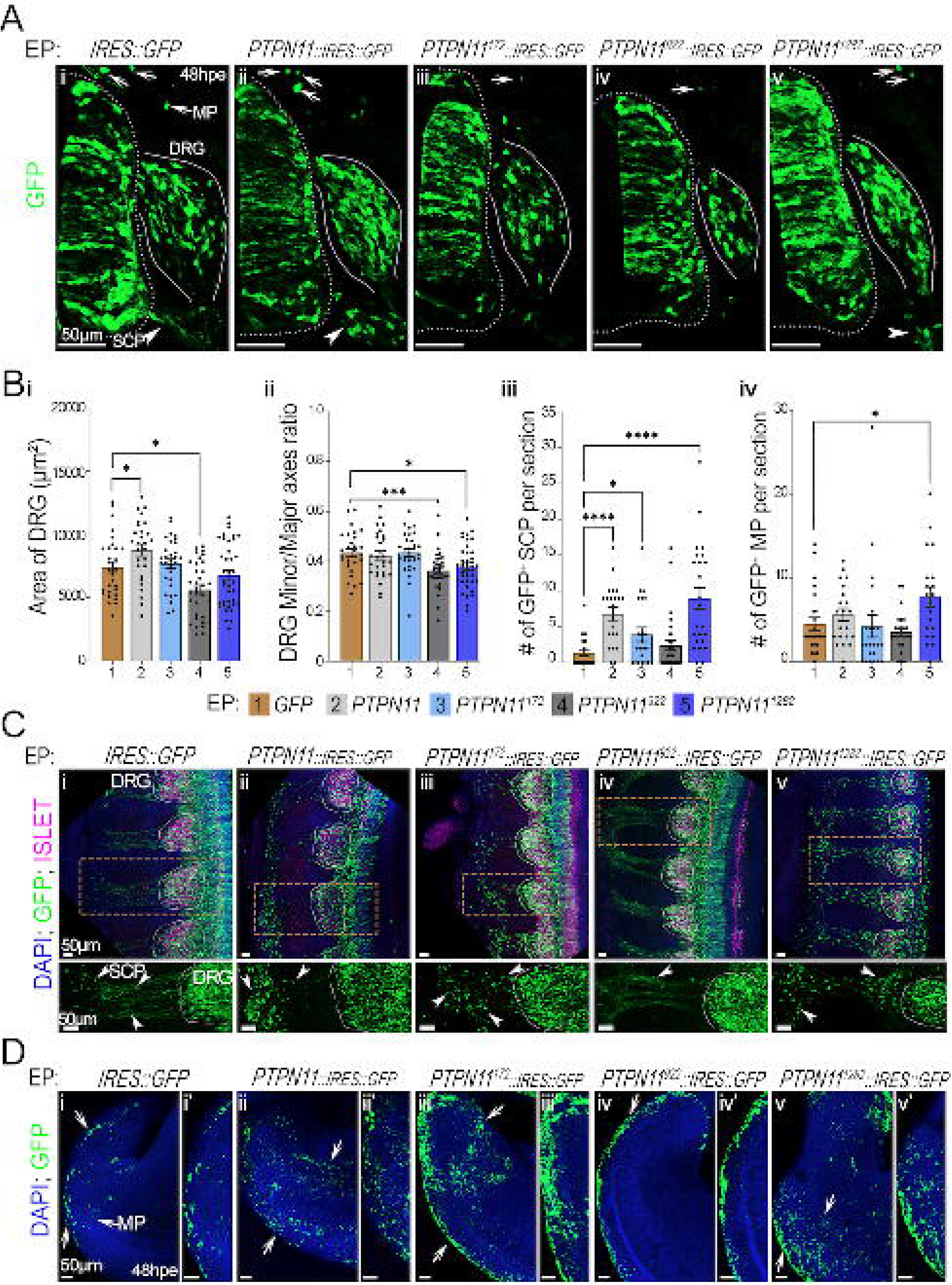
Whole-embryo distribution of *PTPN11* WT- or variant-expressing cells. **(A)** Magnified panels centered on the electroporated side of the sections shown in (Figure 4Ai–v). Immunolabeling GFP (green) on transverse sections of chick embryos at 48 hpe with the indicated constructs. White arrows towards MP, and white arrowheads point at SCP. Scale bars: 50 µm. **(B)** Quantification of DRG area (µm²) (i), the ratio of the minor to major axis of the DRG (ii), the number of SCPs along the ventral root (iii), and the number of MPs under the skin (v) in embryos at 48 hpe with the indicated constructs (n > 6 embryos per condition with one to three sections per embryos). Dots: value per section; bars: mean ± s.e.m. Statistical analysis was performed using unpaired Mann-Whitney tests between the *pCIG* control and each conditions (*P < 0.05, **P < 0.01, ***P < 0.001, ****P < 0.0001). **(C)** Representative Z-projections from the DRG to the ventral root nerve at anterior thoracic levels of whole-mount chick embryos immunolabeled for ISLET1/2 (magenta), GFP (green), and DAPI (blue) on cleared embryos at 48 hpe with the indicated constructs. The bottom panels represent a higher-magnification of the regions indicated by orange dotted boxes in (i–v). Dotted lines delineate the DRG. White arrows: melanocyte precursors (MP), White arrowheads: Schwann cell precursors (SCP) (n > 3 embryos per condition; n = 2 for *PTPN11*^1282^). Scale bars: 50 µm **(D)** Representative Z-projections of the tail skin area of cleared whole-mount chick embryos immunolabeled for GFP (green) and DAPI-stained nuclei at 48 hpe with the indicated constructs. (i′–v′) Higher-magnification panels of the right side of the tail shown in (i–v).

## Discussion

Our integrated approach reveals that *PTPN11* variants span a continuum of functional classes, from strong ERK-dependent hyperactivation to variants with minimal ERK effects, each associated with distinct NCC phenotypes and patient outcomes. First, our analysis of ClinVar and COSMIC datasets shows that pathogenic *PTPN11* mutations cluster in the N-SH2 and PTP domains, likely disrupting SHP2 autoinhibition and catalytic activity. The recurrence of germline and somatic variants in shared regulatory hotspots across NS/NSML and hematologic malignancies, mostly JMML and AML, supports a common mechanism of dysregulation and highlights SHP2’s dual role in development and oncogenesis ^43^. Somatic mutations are enriched at D61, A72, E76, and G507, whereas germline mutations predominantly affect residues N58, G60, D61, E76, F285, T468, G503, and Q510. Both converge on the N-SH2 DE loop (residues 59–62) and the PTP catalytic cleft (residues 460–510), highlighting a shared structural basis for SHP2 dysregulation in development and cancer.

In our pediatric cohort of 18 NS/NSML patients, the ten *PTPN11* variants clustered into four distinct phenotypic classes. The c.1403C>T (T468M) variant was exclusively associated with NSML. All patients presented lentigines, moderate growth impairment and distinctive facial features with preserved cognition. Variants including c.1432A>G (I478V) and c.1282G>A (V428M) were associated with cognitive deficits alongside mild growth and craniofacial involvement, whereas c.1471C>A (P491T) and c.1472C>T (P491L) predominantly affected cardiac and growth phenotypes with limited neurocognitive features. By contrast, variants including c.172A>G (N58D), c.178G>A (G60S), c.844A>G (I282V), c.922A>G (N308D) and c.923A>G (N308S) were associated with more severe, multisystem phenotypes spanning cardiac, growth, cognitive and craniofacial abnormalities. T468M showed context-dependent signaling, consistent with prior reports ^26,28^, highlighting the importance of cellular context. The small number of NSML patients (n=4) limits statistical power, and these findings require validation in larger cohorts. The clinical features support a central role for NCC dysfunction in *PTPN11* disorders including craniofacial dysmorphism and cardiac defects arise from NCC-derived or NCC-influenced tissues ^44^ and pigmentary abnormalities reflect altered melanocyte development ^45^. Additional features, such as sensory neuron involvement and growth impairment, likely result from both NCC-dependent processes and broader systemic effects of dysregulated SHP2 signaling. Consistent with previous animal models showing that NCCs are highly sensitive to both gain- and loss-of-function alterations in SHP2 activity, our chick model identifies NCCs as a particularly vulnerable cell population to *PTPN11* variants. Variants primarily affect NCC allocation and migration rather than lineage identity, suggesting that subtle perturbations in NCC dynamics may underlie the phenotypic variability observed in patients. These populations are also primary targets in neurofibromatosis type 1 (NF1), another RASopathy, highlighting a shared sensitivity of neural crest–derived cells to dysregulation of RAS/MAPK signaling. This convergence is consistent across RASopathies underscores neural crest lineages as a common developmental vulnerability ^30,46^.

In HEK293T cells, where parental HEK293T and MOCK controls displayed low basal SHP2 expression as previously reported ^47,48^, SHP2 expression levels clustered into three groups including WT-like (c.172A>G, c.178G>A, c.1432A>G, c.1471C>A), reduced (c.922A>G), and increased 2- to 5-fold. c.172A>G and c.923A>G drove the strongest ERK hyperactivation (4- to 5-fold), while c.1282G>A showed high SHP2 expression yet failed to increase pERK, indicating paradoxical signaling impairment. The c.922A>G (N308D) variant drove 2.5-fold pERK despite barely detectable protein levels, suggesting that either residual protein below detection is sufficient to hyperactivate MAPK (“catalytic overload”) or the mutation destabilizes autoinhibition, increasing the fraction of active molecules ^22,24^. Variable SHP2 band intensities reflect conformational effects on epitope accessibility rather than abundance differences.

The chick embryo is an accessible *in vivo* embryonic system that harbors both neural progenitors of the spinal cord and NCCs, two lineages prominently affected in RASopathies. Importantly, this system provides a well-characterized framework to study *PTPN11* variants, as modulation of signaling pathways acting upstream and downstream of RAS, including FGF, PI3K, and ERK/MAPK, has been shown to produce defined and reproducible effects on neural tube–derived cell fates, including NCCs-derivatives ^17,19,49^. To determine whether these molecular signatures translate into developmental phenotypes, we expressed three functionally divergent variants, c.172A>G (strong GOF), c.922A>G (low stability, moderate pERK), and c.1282G>A (paradoxical, high SHP2, WT-like pERK), in the chick embryo neural tube. Strikingly, the magnitude of ERK hyperactivation in vitro did not predict the severity of NCC phenotypes in vivo. c.172A>G (N58D), despite driving the strongest pERK, induced only mild NCC effects with a slight increase in SCP population and a modest increase in tail melanocytes, suggesting that trunk NCCs buffer strong GOF effects, or that severe clinical features arise from other cell types. c.922A>G, unlike in the HEK, showed a higher expression in the chick embryos compared to the controls and presents markedly reduced DRG size with an elongated form. While the severe short stature and learning disabilities observed in patients carrying this variant are likely multifactorial, the striking vulnerability of DRG cell population to this variant in the chick model suggests that peripheral nervous system dysfunction, including sensory and autonomic deficits, could contribute to these complex phenotypes. Variant c.1282G>A produced the most dramatic NCC phenotypes (DRG elongation, massive SCP and melanocyte expansion), suggesting that SHP2 may modulate NCC differentiation through ERK-independent mechanisms or via transient/context-specific ERK activation not detected in our steady-state HEK293T assays. Clinically, this variant was associated only with learning disabilities, suggesting that mammalian compensatory mechanisms may attenuate its developmental impact.

### Limitations and Future Directions

The following limitations should be acknowledged. The presence of a co-occurring *ASH1L* variant in a patient with unusually severe or atypical phenotypes suggests potential genetic modifiers, but our cohort size precludes definitive conclusions. The in vitro analyses may not fully recapitulate the signaling context of distinct cell types (e.g., cardiomyocytes, neural crest derivatives, hematopoietic progenitors). The chick embryo model, while powerful for studying NCC migration and differentiation, may not capture all aspects of mammalian development, particularly cardiac and craniofacial morphogenesis. The cohort size (18 patients) limits the statistical power for some genotype-phenotype correlations.

### Conclusions

Our integrated approach combining *in silico* analysis, clinical phenotyping, in vitro signaling, and *in vivo* developmental modeling, reveals that *PTPN11* variants in Noonan spectrum disorders fall into distinct functional classes that cannot be predicted by any single assay. We identify a paradoxical class of variants (exemplified by c.1282G>A, V428M) that drive excessive NCC migration through ERK-independent mechanisms, challenging the prevailing model that NS pathogenesis is primarily driven by MAPK hyperactivation. We also demonstrate lineage-specific sensitivity to SHP2 dosage, with DRG neurons requiring higher SHP2 stability than melanocyte precursors. This work establishes a framework for genotype-guided monitoring and targeted therapeutic strategies in SHP2-associated disorders and underscores the necessity of functional characterization in disease-relevant developmental models for variants of uncertain significance.

## Methods

### Study Cohort and Patient Recruitment

This study included 18 pediatric patients clinically diagnosed with NS or NSML, recruited at the Regional Reference Unit for Advanced Diagnosis of Rare Diseases of Castilla y León (DiERCyL).

All patients were evaluated by clinicians specialized in rare diseases, who performed comprehensive phenotypic assessments. Clinical data were systematically collected using a structured questionnaire specifically designed for the evaluation of NS and NSML features, including growth parameters, craniofacial characteristics, cardiac anomalies, and other associated manifestations. Written informed consent was obtained from all participants or their legal guardians prior to genetic testing. The study was approved by the local Bioethical Committee of the Institute of Biomedical Research of Salamanca.

### DNA Extraction and Next-Generation Sequencing

Peripheral blood samples were collected in EDTA tubes, and genomic DNA was extracted using the MagNA Pure Compact® automated system (Roche Diagnostics), according to the manufacturer’s instructions. DNA concentration and quality were assessed using a BioPhotometer spectrophotometer (Eppendorf). Whole Exome Sequencing (WES) was performed on a NextSeq 550 platform (Illumina). Library preparation was carried out using Truseq DNA library preparation kits (Illumina). Target enrichment was performed using the xGen™ Exome Research Panel (Integrated DNA Technologies, IDT), which captures coding exons and flanking intronic regions. Sequencing reads were aligned to the human reference genome (GRCh37/hg19). Variant calling and annotation were performed using standard bioinformatics pipelines. Candidate variants were validated by Sanger sequencing. Primers were designed using Primer3web, Beacon Designer, and further evaluated for specificity using BLAT and in silico PCR tools from the UCSC Genome Browser. Primers were synthesized by Integrated DNA Technologies (IDT, Coralville, IA, USA).

### Variant Annotation and Pathogenicity Prediction

Variant pathogenicity was assessed according to the American College of Medical Genetics and Genomics (ACMG) guidelines, using VarSome and ClinVar databases, and all classifications were reviewed and confirmed by DiERCyL clinical geneticists. The occurrence of *PTPN11* variants in cancer, as well as available functional evidence, was evaluated using COSMIC and Franklin databases. Population allele frequencies were obtained from the gnomAD database. Evolutionary conservation of variant positions was assessed using PhyloP100 scores retrieved from VarSome. Protein structural analysis was performed using the AlphaFold Protein Structure Database. The predicted structure of SHP2 was downloaded in PDB format and visualized using PyMOL (v3.0) to evaluate the spatial localization of missense variants. Variant distribution across functional domains was analyzed using Mutation Mapper from cBioPortal, based on UniProt domain annotations. Serial Cloner (v2.6.1) was used for *in silico* DNA analysis and cloning design. Potential splicing effects of the ten *PTPN11* variants were assessed using SpliceAI ^50^. Scores range from 0 to 1, and a threshold of ≥0.2 was considered indicative of potential splicing alterations, following established guidelines.

### Cloning Procedures and Plasmid Preparation

The human *PTPN11* coding sequence (encoding SHP2 isoform 1, 593 aa) was obtained from pCMV-SHP2 WT (Addgene #8381). For mammalian expression, *PTPN11* was amplified with SpeI/NotI-tailed primers, subcloned into pBluescript SK(–) (GenBank #X52324.1), and then transferred into the pLVX-IRES-Hyg vector (Clontech #632185). Patient-specific variants were introduced by site-directed mutagenesis using pBSK-*PTPN11* as template. For *in ovo* electroporation, wild-type and mutant *PTPN11* sequences were amplified from pLVX-*PTPN11* and cloned into the XhoI site of pCIG (pCAGGS::IRES::NLS::GFP) using In-Fusion cloning (Takara Bio). Constructs were verified by Sanger sequencing (Nucleus, USAL). Plasmid DNA was prepared using standard kits (Thermo Scientific; Macherey-Nagel) following the manufacturers’ protocols. Primers are listed in Table S1.

### Cell Culture

Both HEK293T (ATCC, #CRL-3216) and NIH/3T3 (ATCC, #CRL-1658) cells were routinely grown in DMEM/F-12 GlutaMAX (Gibco, #A4192001) supplemented with 10% Neo Biotech FBS Advanced (Neo Biotech, #NB-58-0002A) at 37°C in a 5% CO_2_ atmosphere until 70% of confluence was reached. Vectors *pMD2.G* (Addgene, #12259), *psPAX2* (Addgene, #12260) and the pLVX:*PTPN11* constructs were used to produce the lentiviruses in HEK293T cells using transfection reagent PEI (polyethylenimine) (Quimigen, #23966-100). Lentivirus particles were stored at -80°C. HEK293T and NIH/3T3 cells were transduced using polybrene 10 µg/mL (Merck, #TR-1003-G) and selected with hygromycin B 150 µg/mL (Thermo Scientific, #10687010). Cell lines were stored in LN2.

### Western Blotting

Western blot assays were performed to detect SHP2, ERK, AKT, β-actin, phospho-ERK, phospho-AKT Thr308, and phospho-AKT Ser473 proteins (Table S2). Cultured cells were lysed with RIPA buffer (150 mM NaCl, 5mM EDTA, 1% NP-40, 1% sodium deoxycholate, 0.1% SDS, 25 mM Tris-HCl pH 7.6) supplemented with a protease and a phosphatase inhibitor cocktail (Thermo Scientific, #A32955 and #A32957, respectively). After centrifugation at 12,000 rpm for 15 min at 4°C, the lysates were cleared, and the total protein concentration was determined using the Bradford assay (BioRad, #5000006). Protein samples were run on 10% SDS-PAGE gels (acrylamide/bis-acrylamide 29:1) for 70 min at 100 V in 1X SDS-PAGE running buffer. Subsequently, the proteins were transferred onto membranes using Iblot®2 NC Stacks (Invitrogen, #IB23001). Primary antibodies were diluted 1:1000 in TBST with 2.5% BSA, while secondary antibodies were diluted 1:5000 in TBST with 5% milk. Membrane blocking was performed with TBST containing 5% milk for non-phospho antibodies and with TBST containing 5% BSA for phospho-antibodies. After incubation with Pierce ECL Western Blotting Substrate (Thermo Scientific, #32106), membranes were developed using a ChemiDoc MP.

### In Ovo Electroporation of Chicken Embryos

*pCIG-*based constructs (1 µg/µl) were injected into the neural tube of Hamburger and Hamilton (HH) stage 10–11 chick embryos. Electroporation was performed following established protocols ^51^. Embryos were collected at 24- and 48-hours post-electroporation (hpe) and dissected in cold 1× PBS. They were then fixed in 4% paraformaldehyde (PFA) for 1 hour at 4°C. Following fixation, embryos were cryoprotected in 15% sucrose, embedded in gelatin, and cryosectioned at 14 µm. Sections were processed for immunostaining using standard procedures.

### Immunostaining on Histological Sections and Imaging and Image Analyses

Primary antibodies were diluted in PBS containing 1% Triton X-100, 1% BSA, and 0.1% sodium azide (Table S2). Secondary antibodies were used at a dilution of 1:500 in the same buffer. DAPI (1:1000) was included in the secondary antibody solution for nuclear staining. Cryosections were imaged using a Zeiss LSM 980 Airyscan 2 confocal microscope. Image processing was performed using Fiji ^52^. Quantification of SHP2 levels in GFP-positive cells, as well as the percentage of surface area covered by HuC/D-, SOX1-, or SOX10-positive cells within the GFP-positive area, was performed using custom image analysis pipelines ^53^. Cell counts and morphological parameters of dorsal root ganglia (DRG) shape (area, minor and major axis) were quantified using the Cell Counter plugin and region-of-interest (ROI) measurements in Fiji.

### Immunostaining On Cleared Whole Embryos And Imaging

Primary antibodies were diluted in PBS containing 0.5% Triton X-100, 0.1% Tween 20, 1% BSA, and 0.1% sodium azide (Table S2). Secondary antibodies were used at a dilution of 1:500 in the same buffer. DAPI (1:1000) was included in the secondary antibody solution for nuclear staining. Embryos were cleared using the FUnGI optical clearing agent developed in ^54^. Whole embryos were put between two glass slides and were imaged using a Zeiss LSM 980 Airyscan 2 confocal microscope, the Z-stack tool was used for the acquisition. Image processing was performed using Fiji (Schindelin et al., 2019).

### Statistical Analysis

Data normality was assessed using the Shapiro-Wilk test. For normally distributed data, one-way ANOVA followed by Dunnett’s post hoc test was used for multiple comparisons against WT controls. For non-normally distributed data, the Kruskal-Wallis followed by Dunn’s multiple comparisons test was used. For pair-wise comparisons between two groups (in chick embryo experiments), an unpaired, two-tailed Student’s t-test or Mann-Whitney U test was used, depending on the normality of the data. All analyses were performed using GraphPad Prism (version 10.2.0). Data are presented as mean ± SD or SEM from at least three independent experiments or from a minimum of three embryos per condition. For chick data, each analyzed parameter (including DRG area, SCP and MP number), values from individual sections from *n* embryos were pooled, and each dot represents a single measurement from a cut. Statistical significance was defined as follows: *P*<0.05, \**P*<0.01, \*\**P*<0.001, and \*\*\**P*<0.0001.

## Data Availability

All data produced in the present study are available upon reasonable request to the authors.

https://zenodo.org/records/18543932

## Ethics approval and consent to participate

This study was conducted in accordance with the ethical standards of the Declaration of Helsinki and its later amendments. Ethical approval was obtained from the Ethics Committee of the Bioethical Committee. Written informed consent was obtained from all participants (or their legal guardians in the case of minors or vulnerable populations) prior to their inclusion in the study. Participants were informed about the study’s purpose, procedures, potential risks, and their right to withdraw at any time without consequences. All data were anonymized to protect participant confidentiality.

## Availability of data and materials

Due to privacy/ethical restrictions imposed by the Ethics Committee, the raw data supporting this study regarding the patients cannot be made publicly available. However, anonymized data are available from the corresponding author upon reasonable request and with permission from the Ethics Committee. The clincial data that support the findings of this study are available from the corresponding author upon reasonable request. Certain identifying details have been removed to protect participant privacy. The computational pipeline developed by Adiba, S. and used in this study is publicly available on Zenodo at: https://zenodo.org/records/18543932. The rest of the data and materials regarding the experimental work are available upon request.

## Acknowledgements

We gratefully acknowledge support from the FSEEP and GRS for their funding and resources that made this work possible. We also thank UNICAS for their institutional collaboration. Special thanks to all participating families and clinical teams for their invaluable contributions to this research. We sincerely thank the lab technicians for their technical expertise. We also acknowledge the ImagoSeine core facility at the Institut Jacques Monod, a member of France-BioImaging (ANR-10INBS-04) and certified by GIS-IbiSA, especially Thomas Rios for his technical expertise.

## Author contributions

M.R.-M. conducted all experiments and data analyses in cell lines. Chicken embryo experiments and their analyses were carried out jointly by K.C. and M.R.-M., and S.A. designed the image analysis pipelines. K.C. and S.A. jointly quantified chick embryo phenotypes. Conceptualization, investigation, and writing including original draft preparation: M.R.M., J.L., and P.P.M.; writing, review and editing: M.R.M., K.C., V.R., S.A., M.I.G., J.L., and P.P.M.; supervision: J.L.; funding acquisition: P.P.M., M.I.G., J.L., and V.R. All authors have read and agreed to the published version of the manuscript.

## Supplementary Figure Legends

**Figure S1. Pathogenic spectra of germline and somatic *PTPN11* variants. (A)** Histogram of phenotypic spectra with the highest prevalence of *PTPN11* germline mutations annotated in ClinVar as pathogenic or likely pathogenic. **(B)** Histogram of cancer subtypes with the highest prevalence of somatic *PTPN11* pathogenic mutations annotated in COSMIC. RAS: RASopathy-related; NS: Noonan syndrome; NSML: Noonan syndrome with multiple lentigines; MC: metachondromatosis; CV: cardiovascular disorders; MI: male infertility; SS: short stature; ASD: autism spectrum disorder; JMML: juvenile myelomonocytic leukemia; UNC: unclassified; LS: hematopoietic and lymphoid systems; CNS: central nervous system; HN: hematopoietic neoplasm; C: carcinoma; LN: Lymphoid neoplasm; MM: malignant melanoma; G: glioma.

**Figure S2. Early NCC migration and subepidermal mesenchymal colonization upon *PTPN11* WT or variant overexpression in chick neural tube embryos**. Z-projections of the skin and underlying mesenchyme at thoracic levels of cleared whole-mount chick embryos immunolabeled for GFP (green) to mark the MPs under the skin and DAPI-stained nuclei (blue) at 48 hpe with the indicated constructs (n > 3 embryos per condition; n = 2 for *PTPN11¹²*□*²*). Groups of MP are indicated by white arrows. Scale bars: 50 µm.

## Supplementary Table Legends

**Table S1. Primers utilized in this study.** The first primer pair amplifies the *PTPN11* coding sequence with SpeI (5′) and NotI (3′) restriction sites (lowercase) for cloning into pBSK and pLVX. The last primer pair includes XhoI sites and homology arms (bold) for In-Fusion cloning into *pCIG*. All remaining primers introduce site-directed mutations (underlined). Forward (F) and reverse (R) orientations are indicated.

**Table S2. Antibodies utilized in the study.** The antibodies are listed with their host species, conjugation, concentration, supplier and reference.

