## Supplementary material for "An integrated computational, clinical, and functional framework for assessing *PTPN11* (SHP2) variant effects on ERK signaling and neural crest cell behavior in Noonan spectrum disorders": Figure S1

### Slide 1
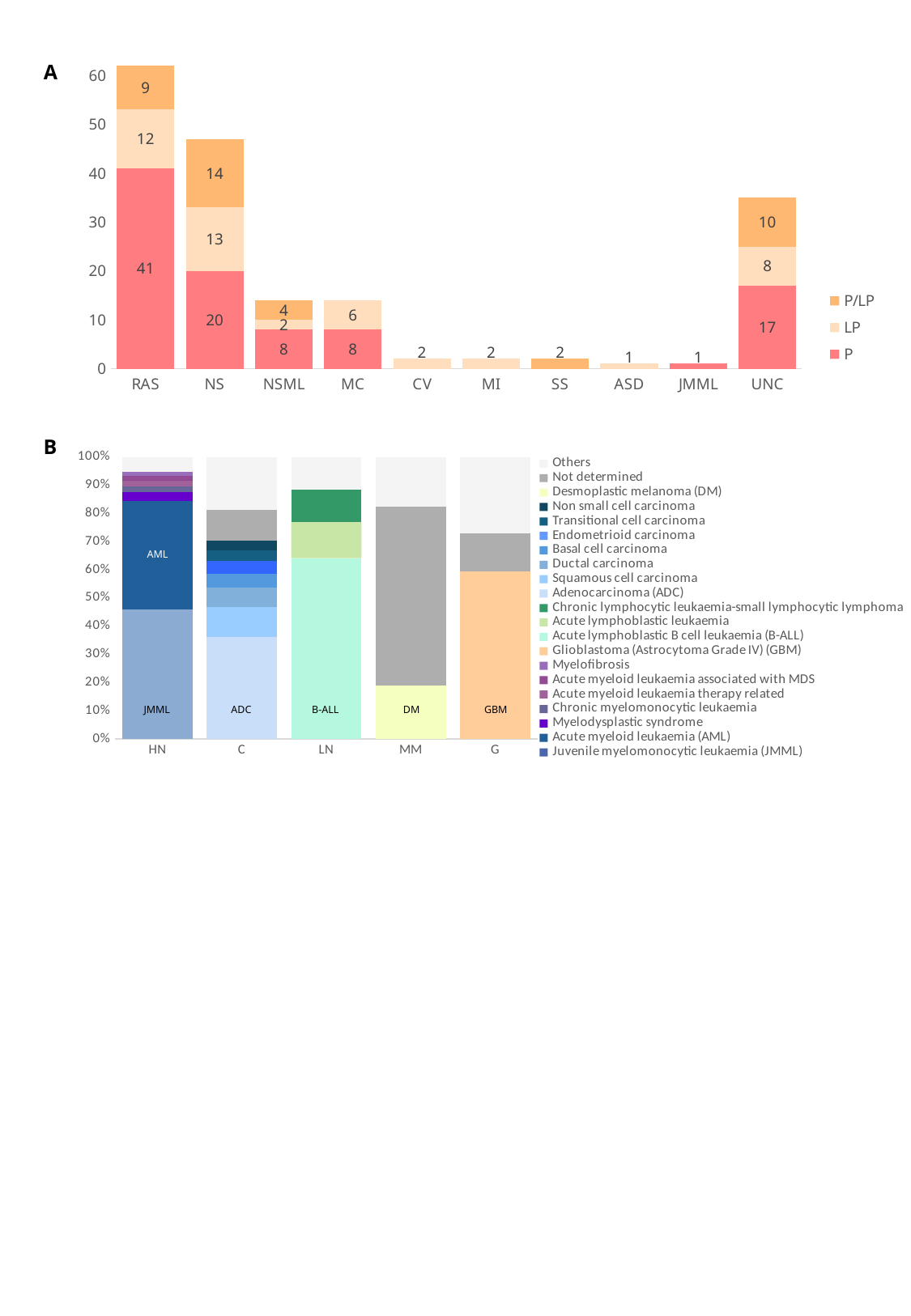

A
#### Chart
| Category | P | LP | P/LP |
|---|---|---|---|
| RAS | 41.0 | 12.0 | 9.0 |
| NS | 20.0 | 13.0 | 14.0 |
| NSML | 8.0 | 2.0 | 4.0 |
| MC | 8.0 | 6.0 | None |
| CV | None | 2.0 | None |
| MI | None | 2.0 | None |
| SS | None | None | 2.0 |
| ASD | None | 1.0 | None |
| JMML | 1.0 | None | None |
| UNC | 17.0 | 8.0 | 10.0 |B
#### Chart
| Category | Juvenile myelomonocytic leukaemia (JMML) | Acute myeloid leukaemia (AML) | Myelodysplastic syndrome | Chronic myelomonocytic leukaemia | Acute myeloid leukaemia therapy related | Acute myeloid leukaemia associated with MDS | Myelofibrosis | Glioblastoma (Astrocytoma Grade IV) (GBM) | Acute lymphoblastic B cell leukaemia (B-ALL) | Acute lymphoblastic leukaemia | Chronic lymphocytic leukaemia-small lymphocytic lymphoma | Adenocarcinoma (ADC) | Squamous cell carcinoma | Ductal carcinoma | Basal cell carcinoma | Endometrioid carcinoma | Transitional cell carcinoma | Non small cell carcinoma | Desmoplastic melanoma (DM) | Not determined | Others |
|---|---|---|---|---|---|---|---|---|---|---|---|---|---|---|---|---|---|---|---|---|---|
| HN | 302.0 | 252.0 | 21.0 | 14.0 | 13.0 | 11.0 | 10.0 | None | None | None | None | None | None | None | None | None | None | None | None | 0.0 | 34.0 |
| C | None | None | None | None | None | None | None | None | None | None | None | 111.0 | 32.0 | 21.0 | 15.0 | 14.0 | 12.0 | 10.0 | None | 34.0 | 57.0 |
| LN | None | None | None | None | None | None | None | None | 129.0 | 26.0 | 23.0 | None | None | None | None | None | None | None | None | 0.0 | 23.0 |
| MM | None | None | None | None | None | None | None | None | None | None | None | None | None | None | None | None | None | None | 12.0 | 40.0 | 11.0 |
| G | None | None | None | None | None | None | None | 35.0 | None | None | None | None | None | None | None | None | None | None | None | 8.0 | 16.0 |AML
JMML
ADC
DM
GBM
B-ALL
