## Supplementary figures and images for "An integrated computational, clinical, and functional framework for assessing *PTPN11* (SHP2) variant effects on ERK signaling and neural crest cell behavior in Noonan spectrum disorders"

### Figure S2

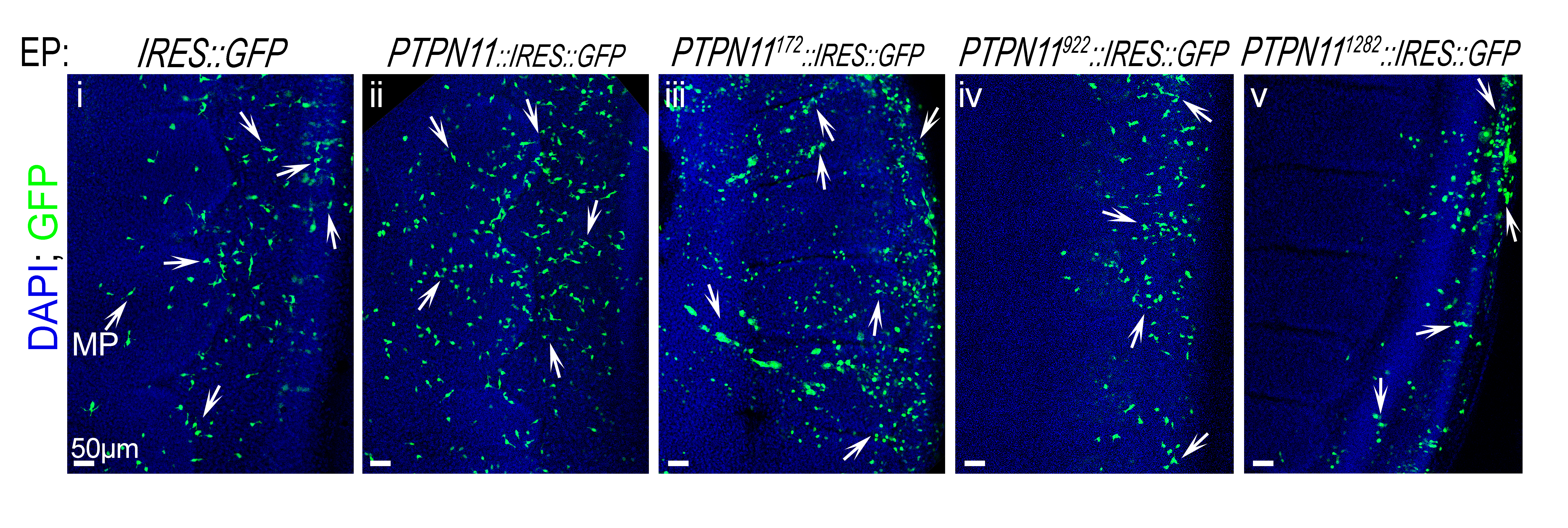
