## Supplementary material for "An integrated computational, clinical, and functional framework for assessing *PTPN11* (SHP2) variant effects on ERK signaling and neural crest cell behavior in Noonan spectrum disorders": Table S1

| **Oligonucleotide** | **Sequence (5’ to 3’)** |
| --- | --- |
| *PTPN11* 5’SpeI | actagtATGACATCGCGGAGATGGT |
| *PTPN11* 3’NotI | gcggccgcTCATCTGAAACTTTTCT |
| *PTPN11* c.172 A>G F | CACATCAAGATTCAGGACACTGGTGATTACT |
| *PTPN11* c.172 A>G R | AGTAATCACCAGTGTCCTGAATCTTGATGTG |
| *PTPN11* c.178 G>A F | CAAGATTCAGAACACTAGTGATTACTATGACCT |
| *PTPN11* c.178 G>A R | AGGTCATAGTAATCACTAGTGTTCTGAATCTTG |
| *PTPN11* c.844 A>G F | AATAGATATAAAAACGTCCTGCCCTTTGATC |
| *PTPN11* c.844 A>G R | GATCAAAGGGCAGGACGTTTTTATATCTATT |
| *PTPN11* c.922 A>G F | CAGATTACATCAATGCAGATATCATCATGCCTG |
| *PTPN11* c.922 A>G R | CAGGCATGATGATATCTGCATTGATGTAATCTG |
| *PTPN11* c.923 A>G F | GATTACATCAATGCAAGTATCATCATGCCTGAA |
| *PTPN11* c.923 A>G R | TTCAGGCATGATGATACTTGCATTGATGTAATC |
| *PTPN11* c.1282 G>A F | TGGCCGGACCACGGCATGCCCAGCGACCCT |
| *PTPN11* c.1282 G>A R | AGGGTCGCTGGGCATGCCGTGGTCCGGCCA |
| *PTPN11* c.1403 C>T F | TTGGCCGGACAGGGATGTTCATTGTGATTGA |
| *PTPN11* c.1403 C>T R | TCAATCACAATGAACATCCCTGTCCGGCCAA |
| *PTPN11* c.1432 A>G F | GATATTCTTATTGACGTCATCAGAGAGAAAGGTG |
| *PTPN11* c.1432 A>G R | CACCTTTCTCTCTGATGACGTCAATAAGAATATC |
| *PTPN11* c.1471 C>A F | TGCGATATTGACGTTACCAAAACCATCCAGA |
| *PTPN11* c.1471 C>A R | TCTGGATGGTTTTGGTAACGTCAATATCGCA |
| *PTPN11* c.1472 C>T F | TGCGATATTGACGTTCTCAAAACCATCCAGATG |
| *PTPN11* c.1472 C>T R | CATCTGGATGGTTTTGAGAACGTCAATATCGCA |
| *PTPN11* 5’XhoI F – pCIG | **CAAAGAATTG**ctcgagATGACATCGCGGAGATGGTT |
| *PTPN11* 3’XhoI R – pCIG | **GTCGATCGAC**ctcgagTCATCTGAAACTTTTCTGCTGTTGC |
