## Supplementary material for "An integrated computational, clinical, and functional framework for assessing *PTPN11* (SHP2) variant effects on ERK signaling and neural crest cell behavior in Noonan spectrum disorders": Table S2

| **Antigens** | | **Species** | **Conjugation** | **Dilution** | **Provider (reference)** | **RRID** |
| --- | --- | --- | --- | --- | --- | --- |
| Actin hFab | | - | Rhodamine | 1/1000 | Bio-RAd #12004164 | - |
| AKT | | Mouse | - | 1/1000 | Santa Cruz #5298 | AB_626658 |
| HuC/D | | Mouse | - | 1/1000 | Thermo Fisher Scientific #16A11 | AB_221448 |
| Chicken | | Donkey | Alexa 488 | 1/500 | Abcam #ab13970 | AB_90755 |
| ERK1/ERK2 | | Rabbit | - | 1/1000 | Invitrogen #44654G | AB_2533710 |
| GFP | | Chicken | - | 1/1000 | Abcam #ab13970 | AB_90755 |
| Isl1 & Isl2 (Islet) | | Mouse | - | 1/50 | DHSB #clone 39.4 | [AB_2314683](http://antibodyregistry.org/AB_2314683) |
| Mouse | | Goat | - | 1/5000 | Invitrogen #G21040 | AB_2536527 |
| Mouse | | Donkey | Alexa 568 | 1/500 | Invitrogen #A10037 | AB_2534116 |
| Phospho-AKT Ser473 | | Mouse | - | 1/1000 | Proteintech #66444-1-Ig | AB_2877033 |
| Phospho-AKT Thr308 | | Rabbit | - | 1/1000 | Proteintech #29163-1-AP | AB_2877777 |
| Phospho-p44/42 ERK1/2 | | Rabbit | - | 1/1000 | Cell Signaling #9101 | AB_331646 |
| Rabbit | | Goat | - | 1/5000 | Invitrogen #G21234 | AB_2536530 |
| Rabbit | | Donkey | Alexa 647 | 1/500 | Invitrogen #A31573 | AB_2536183 |
| SHP2 | | Rabbit | - | WB: 1/1000  IF: 1/500 | Invitrogen #PA520279 | AB_11153412 |
| SOX1 | | Goat | - | 1/200 | R&D systems #AF3369 | - |
| SOX10 | | Goat | - | 1/200 | Santa Cruz #sc-17342 | AB_2195374 |
